# Association between digital biomarkers, loneliness and social isolation: a systematic review and meta-analysis

**DOI:** 10.1101/2025.01.16.25320671

**Authors:** Yolanda Lau, Natalia Chemas, Heema Ajeet Gokani, Rachel Morrell, Harisd Phannarus, Claudia Cooper, Zuzana Walker, Harriet Demnitz-King, Natalie L Marchant

## Abstract

**Question:** What is the current evidence base for the association between digital biomarkers from wrist-worn wearables, loneliness and social isolation in adults?

**Study selection and analysis:** We systematically searched six databases from inception to 24^th^ September, 2024. We narratively synthesised findings and pooled effect sizes using random-effects meta-analyses where possible.

**Findings:** We included 14 studies from 12 articles (12 assessing loneliness, two assessing social isolation). Eight studies used sleep metrics, four used physical activity metrics, and two studies used machine learning approaches. Three meta-analyses were conducted: worse sleep efficiency (SE), but not total sleep time or sleep onset latency, was associated with higher loneliness (Fisher’s z = −0.20, 95% CI −0.34 to −0.06, p = 0.006). Two studies examined wake after sleep onset (WASO), and found longer periods of WASO were associated with higher loneliness. These findings on loneliness were echoed in the study examining social isolation. One study found that lower total physical activity was associated with higher levels of loneliness and social isolation, while other activity intensities showed mixed evidence. Machine learning studies demonstrated high accuracy in predicting loneliness, though models using digital biomarkers from smartphones provided better accuracy.

**Conclusions:** Worse SE, more WASO, and lower total physical activity were associated with loneliness and social isolation, particularly in middle- and older-age. Digital biomarker-based machine learning studies are sparse but show potential in predicting loneliness. Leveraging digital biomarkers as proxy markers of loneliness and social isolation could facilitate early detection of these conditions.

**Key messages of the articles:** *What is already known on this topic:* Loneliness and social isolation are linked to negative health outcomes, including increased dementia risk. Digital biomarkers have shown potential in detecting mental health conditions and symptoms, but no systematic review has explored their association with loneliness and social isolation.

*What this study adds:* This review identified 14 studies examining the association between digital biomarkers and loneliness or social isolation. Worse sleep efficiency, more wake after sleep onset, and lower physical activity were associated with higher levels of loneliness and social isolation.

*How this study might affect research, practice or policy:* Digital biomarkers could support the early detection of loneliness and social isolation, enabling timely intervention.

## INTRODUCTION

Loneliness and social isolation are global public health concerns across all age groups^1,2^, and are particularly prevalent in older adults^3^. Loneliness is a negative emotional state that exists when there is a discrepancy between the desired and perceived actual interpersonal relationships^4^. It is a subjective measure, distinct from social isolation which measures objective social interaction^5^.

Existing methods to measure loneliness and social isolation primarily rely on self-report questionnaires. While these questionnaires are valid measurement methods, they can be influenced by social desirability and recall bias. Questionnaires often capture a general sense of loneliness or social isolation, potentially missing important information as these experiences can fluctuate. For continuous assessment, these measures would need to be administered repeatedly, which would pose additional burden on individuals.

Digital biomarkers are objective, quantifiable, physiological and behavioural data collected using digital devices^6^. They have some benefits over existing assessments of loneliness and social isolation as they can collect objective continuous data. One way to collect digital biomarkers is through wrist-worn wearables (e.g., smartwatches), which are increasingly being used in the general public^7^. Wrist-worn wearables can collect a range of behavioural and physiological data such as physical activity, sleep, and heart rate. Some of these variables have been linked to loneliness and social isolation. For example, systematic reviews found that physical activity^8^ and sleep disturbances^9^ were associated with loneliness, while physical activity was associated with social isolation^10^. If digital biomarkers can serve as indicators of loneliness and social isolation, they could facilitate the early detection of these conditions, which individuals might be hesitant to disclose or may not recognise in themselves.

One of the aims of this review was to examine whether digital biomarkers could eventually be used as a screening tool for dementia risk by monitoring surrogate markers of loneliness and social isolation, as two risk factors for dementia^11–13^. Digital biomarkers can passively collect data, making them particularly suitable for older adults, who may face challenges with retrospective recall or reduced access to clinics for in-person assessments.

To our knowledge, no research has systematically reviewed the existing literature on the association between digital biomarkers and loneliness or social isolation. This systematic review aims to address this gap in the literature.

## METHODS

This review adhered to the guidelines outlined in the Preferred Reporting Items for Systematic Reviews and Meta-Analyses (PRISMA) recommendation^14^ and was registered with PROSPERO (CRD42023409995).

### Search strategy

Five databases (Medline, Embase, PsychINFO, Web of Science, CINAHL) were systematically searched from inception to September 2024. Unpublished grey literature searches were conducted on ProQuest Dissertations and Theses Global. The first 300 papers on Google Scholar were checked as per recommendations^15^. Furthermore, references of eligible papers were checked to identify any eligible studies.

The search strategy employed a combination of two search term strings linked with ‘AND’, each representing a concept related to the research question: (i) digital biomarkers, and (ii) psychosocial risk factors of dementia (see Table S1). The initial search was conducted on 27^th^ April, 2023, and updated on 24^th^ September, 2024.

### Study selection

Two reviewers independently screened titles and abstracts of all the identified studies, followed by full-text screening. Any disagreements were resolved through discussion involving a third reviewer. Inter-rater agreement was evaluated using Cohen’s kappa coefficient.

This review focused on studies that examined loneliness or social isolation, and was embedded within a larger project focusing on psychosocial risk factors of dementia (CRD42023409995). Studies were eligible if they met the following criteria: (i) cross-sectional or longitudinal, (ii), included data from adults aged ≥18 years, (iii) utilised research or consumer grade wrist-worn devices, (iv), evaluated loneliness or social isolation using self-reported standardised assessments (e.g., questionnaire), (v) written in English language, and (vi) examined the association between digital biomarkers obtained from wrist-worn devices and loneliness and/or social isolation. Studies were excluded if (i) they focused on a sample diagnosed with neurodegenerative disorders or physical health conditions, and (ii) they primarily focused on participants with a psychiatric comorbidity, neurological disorders or stroke (>50% of the sample).

### Data extraction and quality assessment

Two independent reviewers extracted all relevant information and assessed the quality of each eligible study, with any disagreements resolved by a third reviewer.

For studies using inferential statistical approaches, the National Heart, Lung and Blood Institute (NHLBI) assessment tool was used^16^. Four criteria that are relevant to longitudinal study designs were deemed inapplicable for cross-sectional studies, and were therefore omitted from assessments of cross-sectional studies^17^. Ratings were averaged, and each study was categorised as “Good” (≥80), “Fair” (50–80%), or “Poor” (<50%).

For studies using machine learning approaches, there is currently no validated quality tool available. The modified Assessment of Diagnostic Accuracy Studies 2 (QUADAS-2) was used^18^. It assesses the risk of bias of studies on four domains (participants, index text [artificial intelligence algorithms], reference standard [ground truth], and analysis), and evaluates applicability across three domains (excluding the analysis domain).

### Synthesis and analysis

A meta-analysis was conducted when two or more standardized effect sizes were available for a digital biomarker type using random-effects meta-analyses for loneliness and social isolation, separately. When standardized effect sizes were unavailable, other available data were used for calculations where possible. All analyses were conducted using the ‘*metafor’* package in R (version 4.2.2).

When multiple effect sizes were reported in a study, the effect size was selected based on an *a priori* determined hierarchy. We prioritised: (i) the largest sample size, and (ii) unadjusted over adjusted estimates, when more than one study used the same cohort. For each meta-analysis, heterogeneity was assessed using *I^2^*. If ten or more studies were included in a meta-analysis, publication bias would be assessed^19^.

A narrative synthesis was conducted, with studies synthesised based on whether loneliness or social isolation was assessed, and the type of digital biomarkers examined. Studies using machine learning techniques were also synthesised qualitatively.

## RESULTS

### Study selection

After removal of duplicates and screening, a total of 14 studies (from 12 articles), were included (Figure. 1). Inter-rater reliability was 0.73 (substantial agreement) for title/abstract, and 0.83 (excellent agreement) for full-text screening.

**Figure 1.**
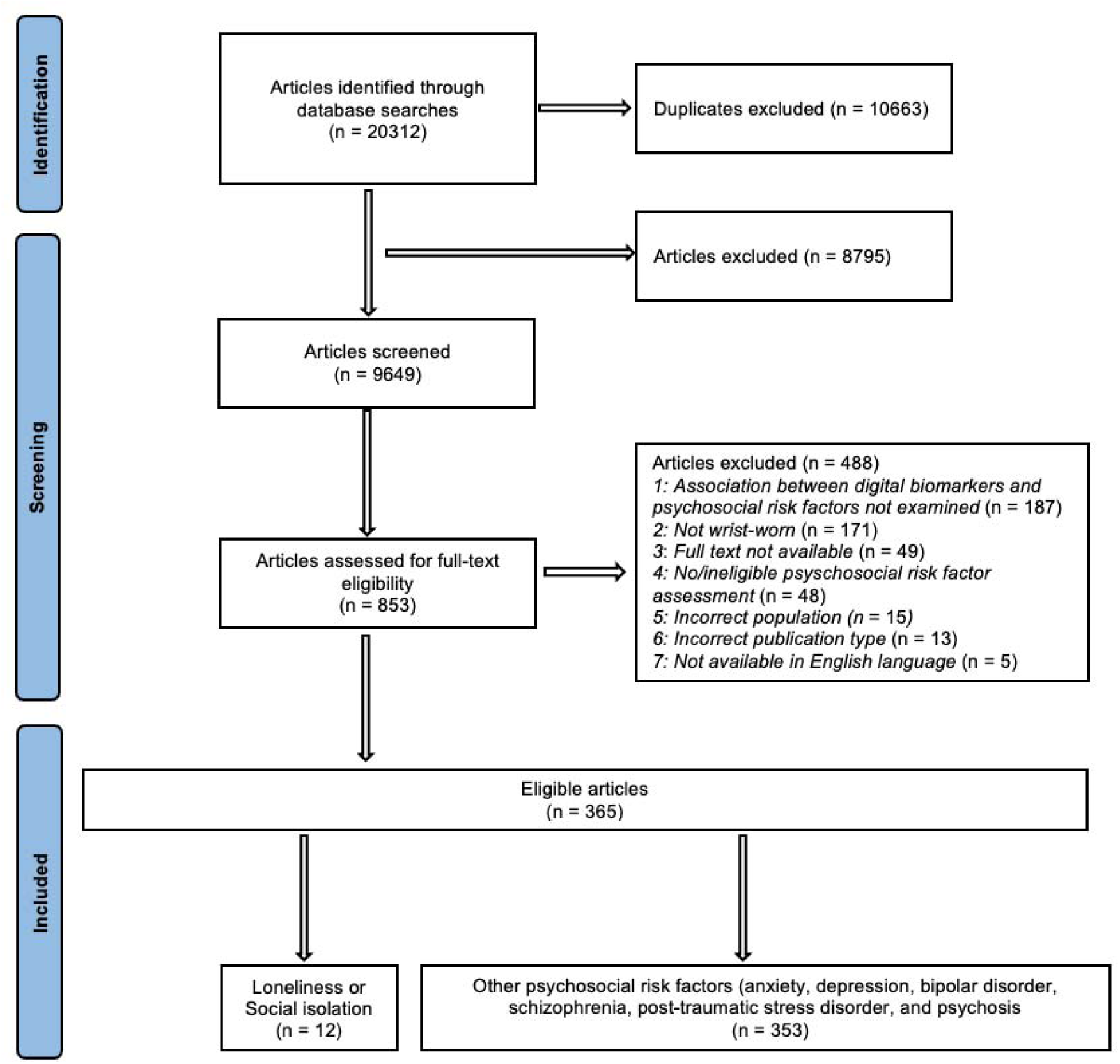
PRISMA flow diagram illustrating the systematic review process

### Study and participants characteristics

Of the 14 studies, 12 (85.7%) used inferential statistical approaches and two (14.3%) used machine learning approaches. Characteristics of the included studies are provided in Table 1, Table 2 and Table S4.

**Table 1.**
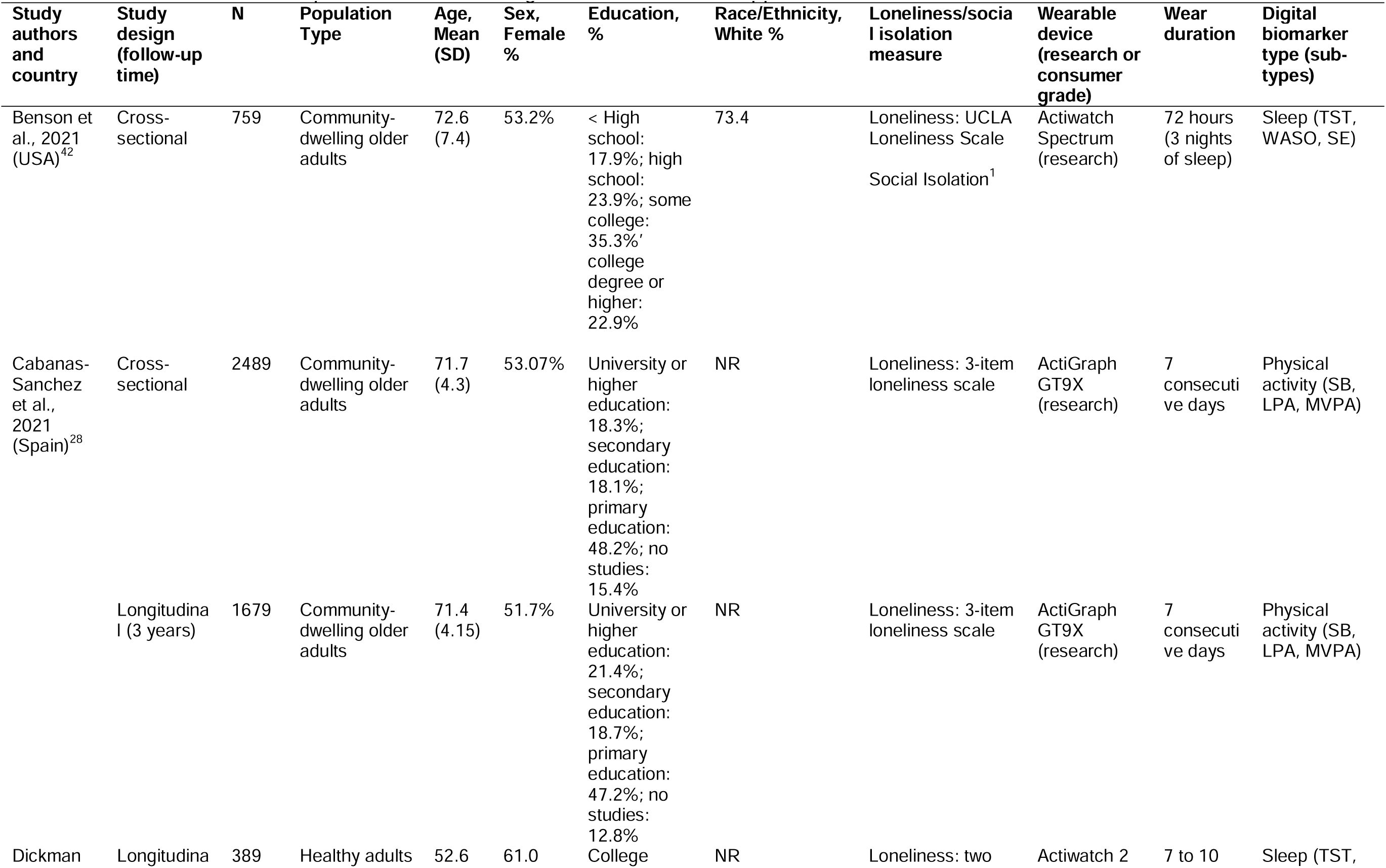
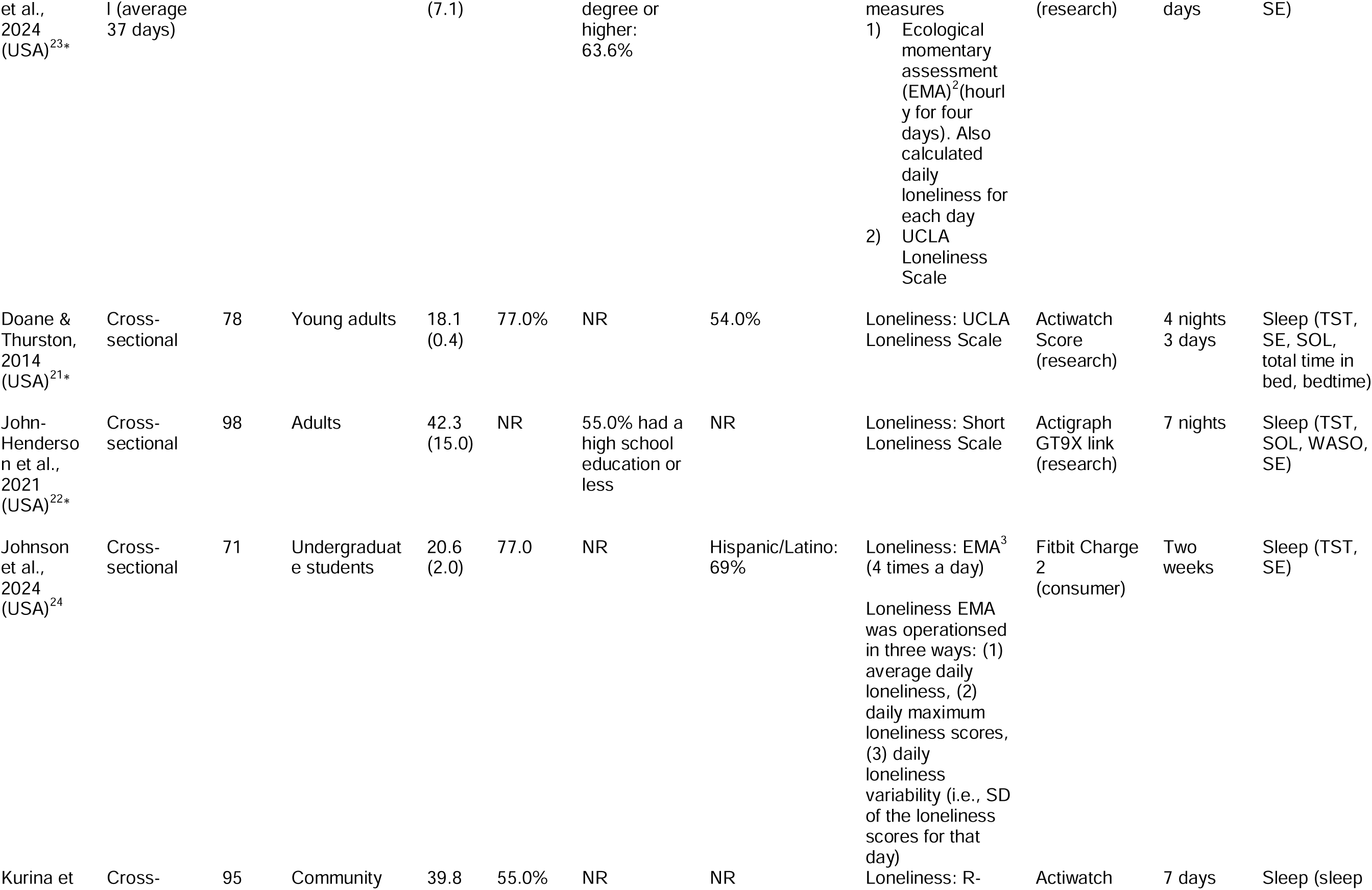
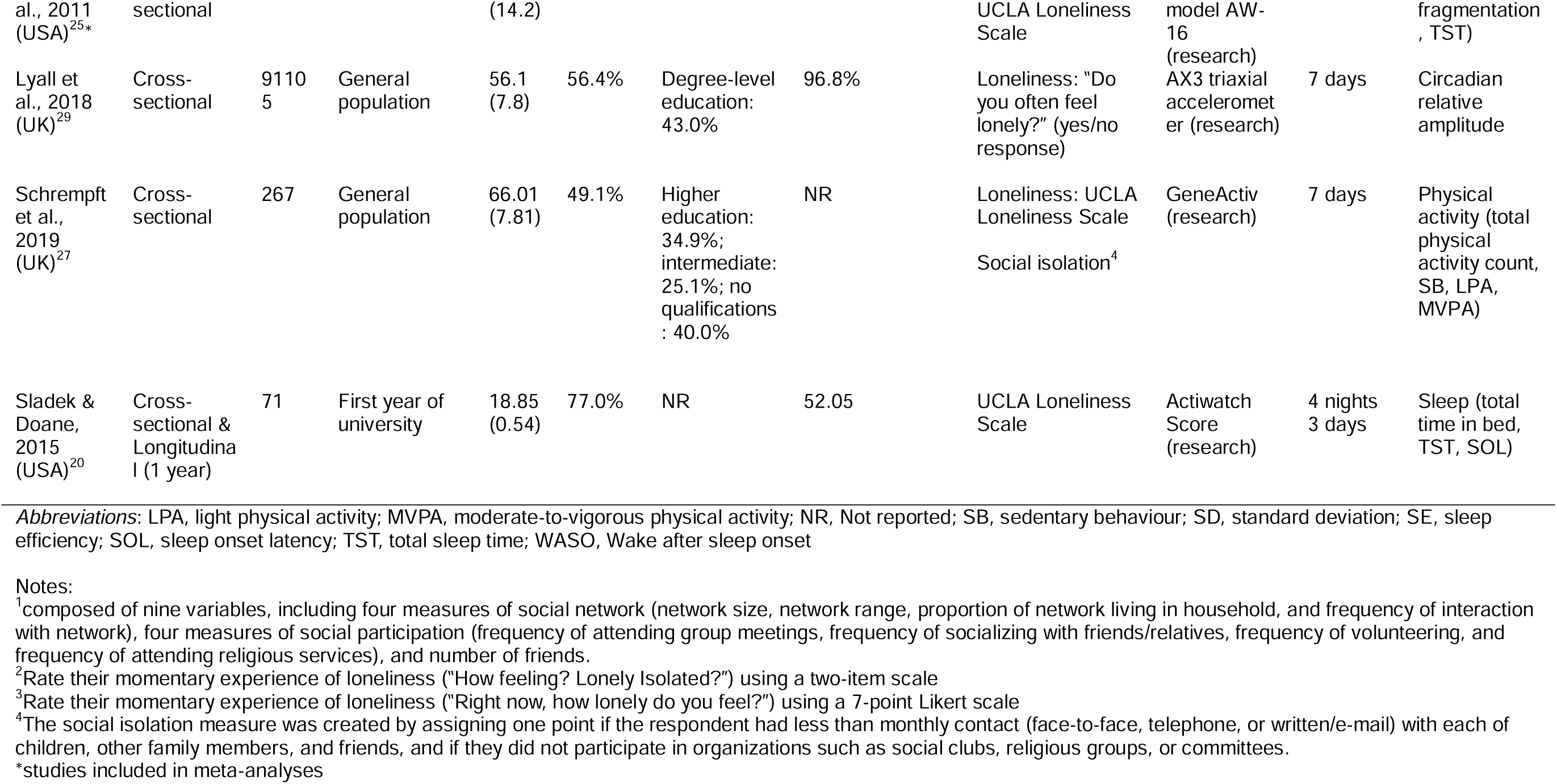
Characteristics of samples from articles using inferential statistical approaches.

**Table 2.**
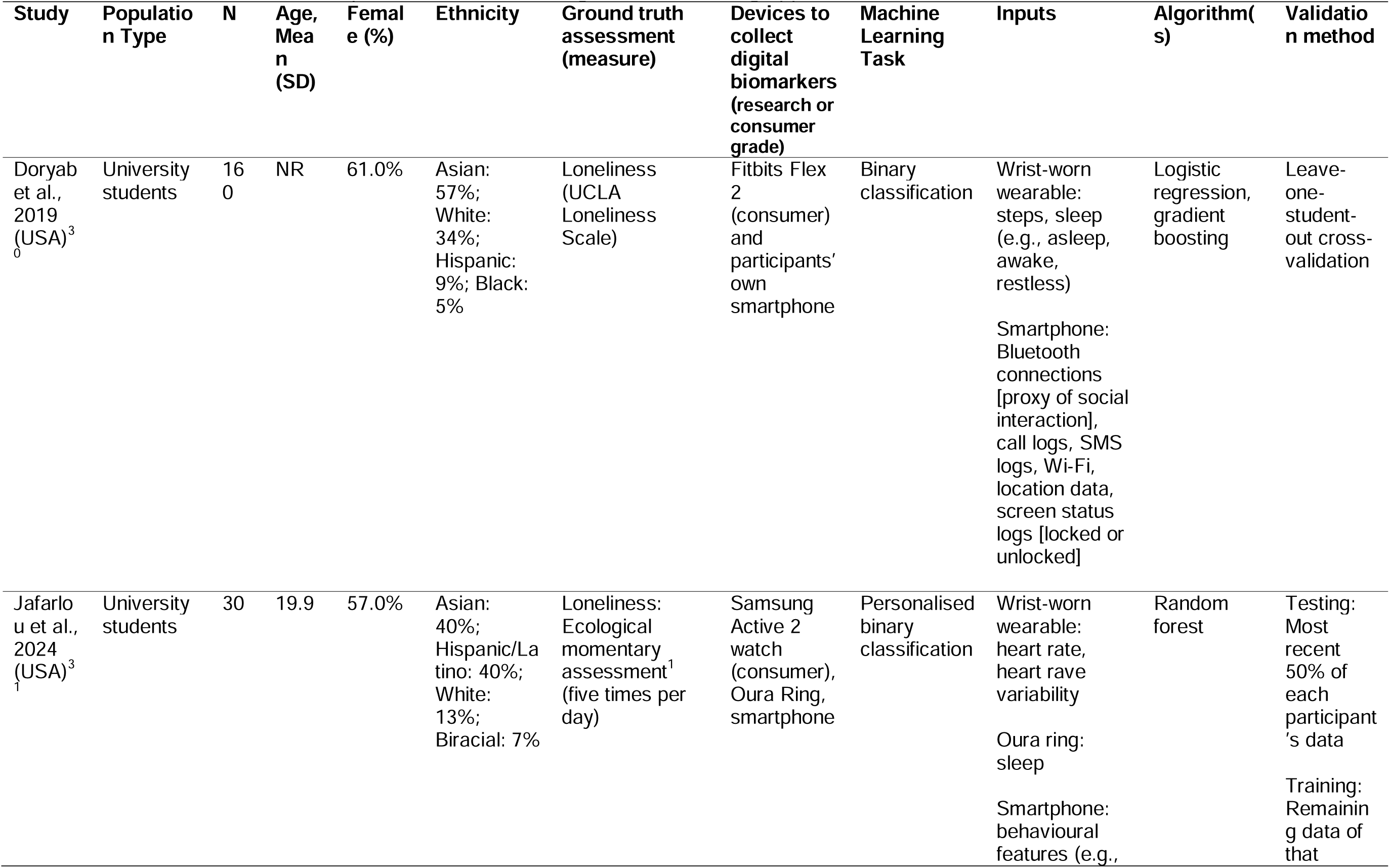
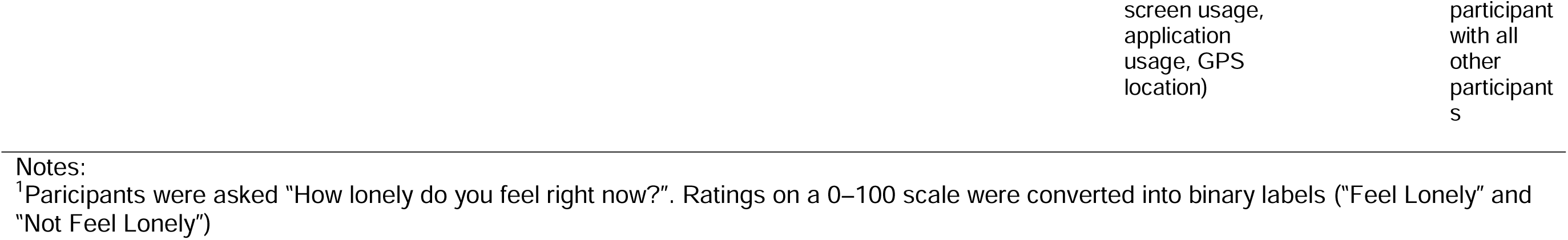
Characteristics of the sample from the article using machine learning approaches.

### Studies using inferential statistical approaches

Twelve studies employed inferential statistical approaches. Of these, ten focused on loneliness, with two additionally examining social isolation. Three studies examined both cross-sectional and longitudinal relationships.

Demographic reporting is based on ten studies because the two social isolation studies were from the same cohorts as the loneliness studies. Sample sizes varied considerably (71 to 91,105 participants; median = 183). Four (40.0%) studies included young adults (18-39 years), three (30.0%) studies included middle-aged adults (40-59 years), and three (30.0%) studies included older adults. Sex composition was reported in nine studies, with a median of 56.4% female participants (49.1% to 77.0%). Six studies (60.0%) reported educational data and five studies (50.0%) reported ethnicity data (Table 1). Most studies were conducted in USA (*k* = 7, 70.0%).

Six different measures were used across the studies that assessed loneliness, with just over half of the articles utilising the UCLA Loneliness Scale (*k* = 6, 60.0%). The two studies assessing social isolation used self-report questionaries with combinations of different variables (e.g., social network, social participation) (Table 1).

All but one study used research-grade wrist-worn wearables (*k* = 11, 91.7%). The majority of articles assessed sleep (k = 8, 66.7%), focusing on metrics such as total sleep time (TST), wake after sleep onset (WASO; the amount of time spent awake after initially falling asleep), sleep onset latency (SOL; the time taken to fall asleep), and sleep efficiency (SE; the ratio of time spent asleep while in bed to the total time spent in bed). Four studies assessed physical activity (33.3%), such as sedentary behaviour, light physical activity, and moderate-to-vigorous physical activity (MVPA).

### Studies using machine learning approaches

Characteristics of the two studies using machine learning approaches to predict loneliness are summarised in Table 2. Both studies focused on university students, and the sample size ranged from 60 to 160.

### Quality assessment

For articles that included both cross-sectional and longitudinal analyses, quality assessment was based on the longitudinal analyses. Of the ten articles using inferential statistical approaches, over half of the studies (*k* = 6, 60.0%) received a ‘Good’ quality rating, the remainder received a ‘Fair’ rating (*k* = 4, 40.0%) (Table S2).

The two machine learning articles showed low risk of bias for reference standard and analysis. For the participants domain, they were rated as either low or high risk. For the index test domain, they were rated as either low risk or unclear risk. There were low concerns regarding applicability for participants, index test, and reference standard (Table S3).

### Studies using inferential statistical approaches: sleep

Results of the included studies are summarised in Table S5.

### Sleep onset latency (SOL)

Three studies examined the relation between SOL and loneliness^20–22^. A meta-analysis revealed no association (k = 2, N = 176; Fisher’s z = 0.09, 95% confidence interval [CI] = −0.13 to 0.31, p = 0.43; Figure 2a), with evidence of high heterogeneity (I² = 51.4%). One study with young adults, not eligible for meta-analysis, also reported no association^20^.

**Figure 2.**
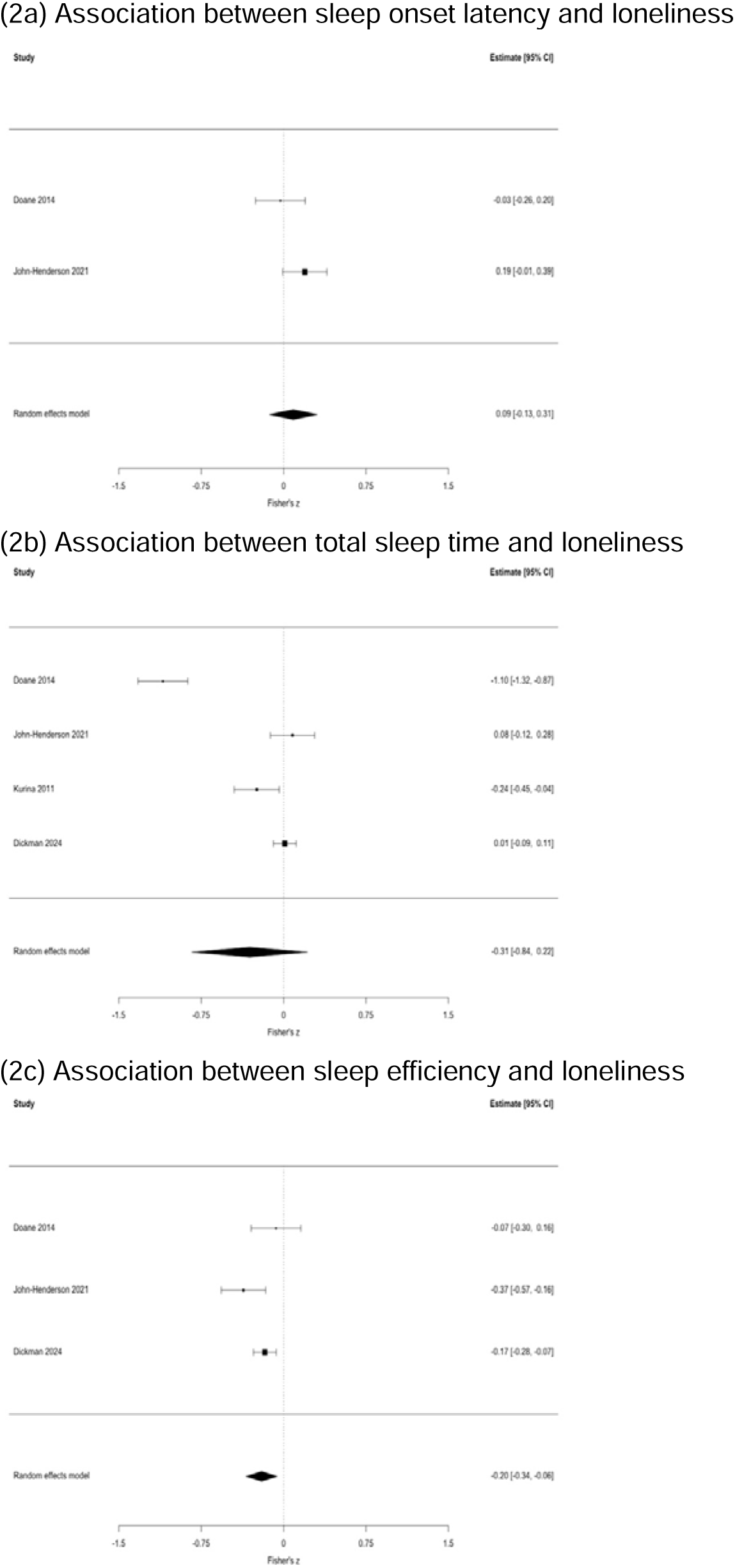
Plots of the associations between sleep metrics and loneliness. Effect sizes are Fisher’s z with corresponding 95% confidence intervals (CI).

### Total sleep time (TST)

The association between TST and loneliness was examined in six studies^20–25^. A meta-analysis revealed no association between TST and loneliness (k = 4, N = 632; Fisher’s z = −0.31, 95% CI = −0.84 to 0.22, p = 0.25; Figure 2b), with evidence of high between-study heterogeneity (I^2^ = 97.2%). Two studies of young adults, not included in the meta-analysis, also found no associations^20,24^. The association between TST and social isolation was examined in one study, no association was observed^26^.

### Sleep efficiency (SE)

The association between SE and loneliness was examined in five studies^21–24,26^. Worse SE was associated with greater levels of loneliness (k = 3, N = 537; Fisher’s z = −0.20, 95% CI = −0.34 to 0-0.06, p = 0.006; Figure 2c), with evidence of high heterogeneity between studies (I^2^ = 51.4%).

Additionally, in two studies not eligible for meta-analysis, the one in older adults reported results that aligned with these findings^26^, while the other in young adults found no association^24^.

The one study that examined social isolation, in older adults, found that worse SE was associated with greater levels of social isolation^26^.

### Wake after sleep onset (WASO)

The association between WASO and loneliness was examined in two studies^22,26^, and the association between WASO and social isolation was examined in one study^26^. All studies were conducted with middle-aged or older adults and found that greater WASO was associated with greater levels of loneliness and social isolation.

### Other sleep metrics

Three studies investigated the association between other sleep metrics and loneliness^20,21,25^. Bedtime and total time in bed were not associated with loneliness^20,21^, but higher levels of loneliness were associated with higher levels of sleep fragmentation (a measure of restlessness during the sleep period)^25^.

### Studies using inferential statistical approaches: physical activity

Two studies investigated the cross-sectional association between physical activity and loneliness in older adults^27,28^. In both studies, loneliness was not associated with sedentary behaviour or light physical activity. Regarding MVPA, one study found no association with loneliness^27^, however the other study found that replacing sedentary behaviour, or light physical activity with MVPA was associated with lower loneliness^28^. Examining physical activity at any intensity, total physical activity count was associated with lower loneliness^27^, however no associations were observed between baseline physical activity (sedentary behaviour, light physical activity, or MVPA) and change in loneliness at 3-year follow-up^28^.

Another study in middle aged adults examined the association between circadian relative amplitude – the difference between the most active continuous 10-hour period and the least active continuous 5-hour period in 24 hours, with higher levels indicating a clearer distinction between the most and least active periods of the day. They found that that lower circadian relative amplitude was associated with higher odds of reporting loneliness^29^.

In the only study that investigated the association between physical activity and social isolation, more total physical activity count was associated with lower levels of social isolation in older adults, echoing the finding with loneliness^27^. Further, lower sedentary behaviour, more light physical activity and more MVPA were associated with lower levels of social isolation.

### Studies using machine learning approaches

#### Results of the two studies are summarised in Table S6

One study used data collected from wrist-worn devices (steps, sleep status) and smartphones to predict loneliness in young adults^30^. They found that the models using steps, sleep (metrics not specified), call logs, location patterns, and screen status achieved the highest accuracy (80.2%) in detecting high levels of loneliness. They also found the model achieved 88.4% accuracy in predicting changes in loneliness levels over a school semester, using Bluetooth, call logs, and location maps, but this final model did not include any data from wrist-worn devices.

Another study used data from wrist-worn wearables (heart rate), smart rings, and smartphones in young adults^31^. They aggregated results from 30 different personal machine learning models. They trained four models: data from (1) smart rings, (2) wrist-worn wearables, (3) smartphones, and (4) all three devices. They found that the model trained on only wrist-worn wearables data achieved the second-best accuracy (78.1%), while the highest accuracy (81.0%) was achieved by the model using data from smartphones. In addition, when considering all the digital biomarkers from all devices, the top six most important features for each participant did not include any data from the wrist-worn wearables.

## DISCUSSION

This systematic review and meta-analysis sought to synthesise evidence regarding the association between digital biomarkers derived from wrist-worn wearables and loneliness and social isolation. Twelve studies using inferential statistical approaches were included, 10 assessed loneliness and two assessed social isolation. Meta-analyses conducted to examine associations between three sleep metrics (SE, TST, SOL) and loneliness revealed that only worse SE was associated with higher loneliness. Our narrative synthesis found that more WASO was associated with higher loneliness and social isolation in middle-aged and older adults. Further, lower physical activity was associated with higher levels of loneliness and social isolation in middle-aged and older adults, while findings regarding different physical intensities were mixed.

### Sleep

Our review found that worse SE and more WASO, but not TST and SOL, were associated with higher levels of loneliness, echoing the findings from our recent meta-analysis on digital biomarkers and anxiety^32^. Moreover, these findings align with previous studies using the Pittsburgh Sleep Quality Index (PSQI) total sore, which found associations between worse subjective sleep and greater levels of loneliness and social isolation in young^33^ and older adults^34–37^.

Looking at specific sleep components of the PSQI, some studies found that longer subjective SOL was associated with higher loneliness levels^33,34,36^. In contrast, our review did not observe associations between SOL measured from wrist-worn devices and loneliness.

However, only one of the three studies in our review asked middle-aged participants to indicate their sleep start time for SOL calculation^22^, without this information, the estimated sleep start time and thus SOL might be less accurate.

Existing studies using PSQI found mixed evidence regarding the association between TST and loneliness^33,34,36,38^. Aligning with our review, a recent meta-analysis found no association between TST and loneliness in adults^39^. Consistent with the findings in our review, a study using polysomnography (gold standard for measuring sleep) found that lonely young adults had worse SE and more WASO than those who were not lonely, but no associations were observed with TST^40^.

Only one study in our review investigated the association between sleep and social isolation and reported that social isolation was associated with worse SE and more WASO, but not with TST, in older adults^26^. Our findings are consistent with existing research that used subjective measures, such as the PSQI total score^35,41^, and subjective sleep duration^42^.

Taken together, these findings suggest that disruptions during sleep (as indicated by worse SE and more WASO) may be more associated with loneliness and social isolation, than the duration or timing of sleep (TST, SOL).

### Physical activity

Our review found that higher total physical activity was associated with lower levels of loneliness and social isolation. However, no or mixed evidence was observed for other physical activity metrics including specific physical activity intensities and loneliness.

Although these findings are based on a very limited number of studies they generally align with the established negative association between self-reported physical activity (assessing different aspects of physical activity, e.g., general physical activity level, specific types of exercises) and loneliness in adults and older adults^43,44^.

Our review included only one study that examined the association between physical activity and social isolation, which found that more total physical activity, light physical activity, and MVPA, and lower sedentary behaviour were associated with lower levels of social isolation in older adults^27^. These findings are consistent with existing research using self-report measures of physical activity which report that higher levels of physical activity were associated with lower social isolation in older adults^45,46^. These findings suggest that engaging in some physical activity is beneficial for reducing loneliness and loneliness, regardless of the intensities of the physical activity.

### Studies using machine learning approaches

Machine learning approaches offer several advantages over inferential statistics because they can incorporate multiple predictors to uncover complex patterns, resulting in potentially better predictive accuracy. The two studies using machine learning techniques found promising results (accuracy: 78.1%-88.4%), comparable to machine learning models using wearable data to predict depression^18^ and anxiety^47^. The first study in our review found that the model without wrist-worn wearable data performed better than models with these data, highlighting the potential importance of behavioural features collected from smartphones^30^. Features such as call logs, which are not captured by wrist-worn wearables, may reflect social connections and interaction, which are related to both loneliness and social isolation. Similarly, the second study did not identify any data from wrist-worn wearables as the top predictors for each participant (the top predictors were behavioural markers from smartphones [e.g., notifications] and heart rate variability features from smart rings), even though the model using only wrist-worn wearable data achieved the second-best accuracy^31^.

However, both studies had small sample sizes, focused on young-adult student populations, and included limited digital biomarkers from the wrist-worn wearables. This limits the generalisability of the findings to other age groups, like older adults, where we observed different patterns in the association between loneliness and sleep and physical activity in younger compared to older populations.

### Older adults

Five studies in our review focused on older adults^26–28^, with evidence that sleep (WASO and SE) and physical activity (total physical activity count and MVPA) are associated with loneliness and social isolation. Although our review only identified a small number of studies in older adults, these findings underscore the potential clinical utility of digital biomarkers in detecting loneliness and social isolation (and dementia risk) among older adults, which could allow early identification and intervention. However, more research using digital biomarkers is needed in this population.

### Limitations and future directions

To our knowledge, this is the first review to examine the association between digital biomarkers and loneliness and social isolation. Key strengths of this review include the rigorous methodological approach (e.g. comprehensive database searches, double screening of all articles). Further, most studies were of good quality, which enhances the validity and reliability of our findings.

This review also has several limitations. First, there were a paucity of studies that measured either social isolation or physical activity; therefore, more research is needed before any firm conclusions can be drawn. Second, the majority of the studies utilised research-grade wrist-worn devices (i.e., devices designed for research purposes). Since the general public only has access to consumer-grade devices (i.e., devices that are available for purchase directly by consumers), future studies first need to examine whether the associations observed between digital biomarkers and loneliness and social isolation are also observed in consumer-grade devices. Third, the majority of the included studies were cross-sectional, limiting the ability to examine the trajectories over time. Finally, more research using machine learning techniques is needed, especially in older adults as loneliness and social isolation are associated with increased dementia risk. Machine learning models using digital biomarkers as predictors, could potentially be used as a screening tool to assist clinicians in identifying individuals experiencing loneliness and social isolation more effectively.

## Conclusion

This systematic review and meta-analysis found that more wake after sleep onset and worse sleep efficiency were associated with higher loneliness and social isolation levels. Although the number of studies looking at physical metrics was limited, there was evidence that more physical activity is associated with lower loneliness and social isolation in middle-aged and older adults. Machine learning approaches showed promise in predicting loneliness, although studies only focused on young adults. Finally, given the high prevalence of loneliness and social isolation in older adults and their association with increased dementia risk, more research is needed to explore the potential of digital biomarkers for the early detection of loneliness, social isolation and dementia risk.

## Data Availability

All data produced in the present study are available upon reasonable request to the authors

## Acknowledgements

Yolanda Lau is a PhD student funded by the Economic & Social Research Council’s London (UBEL) Doctoral Training Partnership (ES/P000592/1), embedded within the APPLE-TREE programme which was funded by an Economic and Social Research Council/National Institute for Health Research programme grant (ES/S010408/1). The views expressed in this publication are those of the author(s) and not necessarily those of the National Institute for Health Research or the Department of Health and Social Care.

## Abbreviations

MVPA: moderate-vigorous physical activity

NHLBI: National Heart, Lung and Blood Institute assessment tool

PRISMA: Preferred Reporting Items for Systematic Reviews and Meta-Analyses

PSQI: Pittsburgh Sleep Quality Index

QUADAS-2: Assessment of Diagnostic Accuracy Studies 2

SE: sleep efficiency

SOL: sleep onset latency

TST: total sleep time

WASO: wake after sleep onset

