## Supplementary materials for "Association between digital biomarkers, loneliness and social isolation: a systematic review and meta-analysis"

**Table S1**. Overview of search terms

**Table S2**. Risk of bias for articles using inferential statistical approaches

**Table S3**. Risk of bias for articles using machine learning approaches

**Table S4**. Summary of sample demographics from studies using inferential statistical approaches according to analysis

**Table S5**. Summary of main results from articles using inferential statistical approaches

**Table S6**. Findings of the articles using machine learning approaches

**Table S1**. Overview of search terms

|  | Search terms^1^ |
| --- | --- |
| Digital biomarkers | (wearable* or "digital biomarker*" or smartwatch* or "smart watch" or "apple watch" or applewatch or fitbit or Samsung or garmin or lg or Huawei or sony or xiaomi or adidas or nike or axivity or pedometer* or "fitness track*" or "fitness monitor*" or acceleromet* or "activity monitor*" or "activity track*" or actigraphy).ti,ab,kw |
| Psychosocial dementia risk factors | (depress* or anxiet* or anxious* or "post-traumatic stress disorder" or PTSD or lonel* or "social isolation" or psychosis or psychotic or schizophreni* or bipolar).ti,ab,kw |

*Abbreviations*: ab = abstract; kw = keyword; ti = title; * = truncation

*Notes*: ^1^Search terms were adapted for each database.

**Table S2**. Risk of bias for articles using inferential statistical approaches

| **Criteria** | **Yes** | **No** | **Other**  **(CD, NR, NA)** |
| --- | --- | --- | --- |
| 1. Was the research question or objective in this paper clearly stated? | 10 (100%) | 0 (0%) | 0 (0%) |
| 2. Was the study population clearly specified and defined? | 10 (100%) | 0 (0%) | 0 (0%) |
| 3. Was the participation rate of eligible persons at least 50%? | 9 (90%) | 1 (10%) | 0 (0%) |
| 4. Were all the subjects selected or recruited from the same or similar populations (including the same time period)? Were inclusion and exclusion criteria for being in the study prespecified and applied uniformly to all participants? | 10 (100%) | 0 (0%) | 0 (0%) |
| 5. Was a sample size justification, power description, or variance and effect estimates provided? | 0 (100%) | 10 (100%) | 0 (0%) |
| 6. For the analyses in this paper, were the exposure(s) of interest measured prior to the outcome(s) being measured? | 2 (20%) | 0 (0%) | 8 (80%) |
| 7. Was the timeframe sufficient so that one could reasonably expect to see an association between exposure and outcome if it existed? | 3 (3%) | 0 (0%) | 7 (70%) |
| 8. For exposures that can vary in amount or level, did the study examine different levels of the exposure as related to the outcome (e.g., categories of exposure, or exposure measured as continuous variable)? | 10 (100%) | 0 (0%) | 0 (0%) |
| 9. Were the exposure measures (independent variables) clearly defined, valid, reliable, and implemented consistently across all study participants? | 10 (100%) | 0 (0%) | 0 (0%) |
| 10. Was the exposure(s) assessed more than once over time? | 3 (3%) | 0 (0%) | 7 (70%) |
| 11. Were the outcome measures (dependent variables) clearly defined, valid, reliable, and implemented consistently across all study participants? | 10 (100%) | 0 (0%) | 0 (0%) |
| 12. Were the outcome assessors blinded to the exposure status of participants? | 0 (0%) | 0 (0%) | 10 (100%) |
| 13. Was loss to follow-up after baseline 20% or less? | 2 (20%) | 0 (0%) | 8 (80%) |
| 14. Were key potential confounding variables measured and adjusted statistically for their impact on the relationship between exposure(s) and outcome(s)? | 9 (90%) | 1 (10%) | 0 (0%) |

*Abbreviations:* CD, cannot determine; NA, not applicable; NR, not reported

*Notes:* National Heart, Lung and Blood Institute (NHLBI) assessment tool was used. Data presented as *k* (%). Criteria 6, 7, 10, and 13 were excluded for cross-sectional studies.

**Table S3**. Risk of bias for articles using machine learning approaches

|  | **Risk of bias** | | | | **Applicability concerns** | | |
| --- | --- | --- | --- | --- | --- | --- | --- |
|  | **Participants** | **Index test** | **Reference standard** | **Analysis** | **Participants** | **Index test** | **Reference standard** |
| **Doryab et al., 2019** | Low risk | Unclear | Low risk | Low risk | Low concerns | Low concerns | Low concerns |
| **Jafarlou et al., 2024** | High risk | Low risk | Low risk | Low risk | Low concerns | Low concerns | Low concerns |

**Table S4**. Summary of sample demographics from studies using inferential statistical approaches according to analysis

|  | **Included in systematic review (*k* = 12)** | | **Loneliness (*k* = 10)** | | **Social Isolation (*k* = 2)** | |
| --- | --- | --- | --- | --- | --- | --- |
|  | *k* | N (%) or Median (range) | *k* | N (%) or Median (range) | *k* | N (%) or Median (range) |
| Study design | 12 |  | 10 |  | 2 |  |
| Cross-sectional |  | 9 (75.0%) |  | 7 (70. 0%) |  | 2 (100%) |
| Cross-sectional & longitudinal |  | 3 (25.0%) |  | 3 (30.0%) |  | 0 (0%) |
| Sample size | 12 | 267 (71-91105) | 10 | 183 (71-91,105) | 2 | 513 (267-759) |
| Age, years | 12 | 66.0 (18.9-72.7) | 10 | 47.4 (18.9-72.7) | 2 | 69.3 (66.0-72.6) |
| Age group | 12 |  | 10 |  | 2 |  |
| Young adults, <40 |  | 4 (33.3%) |  | 4 (40.0%) |  | 0 (0%) |
| Middle-aged adults, 40-59 |  | 3 (25.0%) |  | 3 (30.0%) |  | 0 (0%) |
| Older adults, ≥60 |  | 5 (41.7%) |  | 3 (30.0%) |  | 2 (100%) |
| Sex, female | 11 | 55.0% (49.1%-77.0%) | 9 | 56.4% (49.1-77.0%) | 2 | 51.1% (49.1%-53.2%) |
| Digital biomarker type | 12 |  | 10 |  | 2 |  |
| Sleep |  | 8 (66.7%) |  | 7 (70.0%) |  | 1 (50%) |
| Activity |  | 3 (25.0%) |  | 2 (20.0 %) |  | 1 (50%) |
| Sleep & Activity |  | 1 (8.3%) |  | 1 (10.0 %) |  | 0 (0%) |
| Device type, research-grade | 12 | 11 (91.2%) | 10 | 9 (90.0%) | 2 | 2 (100%) |

**Table S5**. Summary of main results from articles using inferential statistical approaches

| **Study** | **Predictor** | **Outcome** | **Covariates** | **Brief summary** |
| --- | --- | --- | --- | --- |
| Benson et al., 2021 (USA)^32^ | Loneliness | Sleep | Age, sex, education, and race/ethnicity | Loneliness positively associated with WASO and negatively associated with SE. No association observed with TST. |
|  | Social isolation | Sleep | Age, sex, education, and race/ethnicity | Social isolation positively associated with WASO and negatively with SE. No association observed with TST. |
| Cabanas-Sanchez et al., 2021 (Spain)^19^ | Cross-sectional analyses: Sleep or activity | Cross-sectional analyses: Loneliness | Cross-sectional analyses: Sex, and age, educational level, marital status, household economy, smoking status, alcohol consume, total energy intake, BMI, cognitive function, physical function, and chronic diseases | Cross-sectional analyses – isotemporal substitution analyses: Replacing sleep, SB and LPA with MVPA was associated with lower loneliness scores. |
|  | Longitudinal analyses: Baseline sleep and activity | Longitudinal analyses: Follow-up loneliness | Longitudinal analyses: further adjust for predictors at baseline | Longitudinal analyses - No associations observed between replacing sleep with activity and loneliness. |
| Dickman et al., 2024 (USA)^13^ | None | None | None | Greater average loneliness in daily life (measured using Ecological Momentary Assessment) was negatively associated with SE, but not associated with loneliness measured using the UCLA. No association observed with TST. |
| Doane & Thurston, 2014 (USA)^16^ | None | None | None | No associations observed between TST, SOL, total time in bed or bedtime and loneliness. |
| John-Henderson et al., 2021 (USA)^15^ | Correlations: None  Generalised linear mixed models: loneliness | Correlations: None  Generalised linear mixed models: sleep | Correlations: None  Generalised linear mixed models: age, gender, income, anxiety symptoms, depressive symptoms, and adverse childhood experiences | Correlations: loneliness positively associated with WASO and negatively associated SE. No associations observed with TST or SOL.  Generalised linear mixed models: Participants with higher levels of loneliness associated with higher WASO, higher SOL, and lower SE than those with lower levels of loneliness. No association observed with TST. |
| Johnson et al., 2024 (USA)^12^ | Loneliness | Sleep | Race, ethnicity, age, gender, caffeine, nicotine, and alcohol use | No association observed with SE and TST. |
| Kurina et al., 2011 (USA)^17^ | Loneliness | Sleep | Age, sex, body mass index, risk of sleep apnea, and negative affect | Regression: Loneliness associated with more sleep fragmentation and lower TST  Mixed linear models: Loneliness was associated with increase in sleep fragmentation. No association observed with TST. |
| Lyall et al., 2018 (UK)^20^ | Circadian relative amplitude | Loneliness (cases vs. controls) | Age, season at which accelerometry started, sex, ethnicity, and Townsend deprivation score, alcohol intake frequency, smoking status, degree, overall mean acceleration, and body-mass index, and childhood trauma | Lower relative amplitude associated with higher odds of reporting loneliness. |
| Schrempft et al., 2019 (UK)^18^ | Loneliness | Activity | Gender, age, educational attainment, non-pension wealth, marital status, smoking, alcohol consumption, limiting illness, mobility impairment, and self-rated health, depressive symptoms, social isolation | Loneliness negatively associated with total activity count (no longer significant after adjusting for covariates). No associations observed with SB, LPA, or MVPA. |
| Sladek & Doane, 2015 (USA)^11^ | Social Isolation | Activity | Gender, age, educational attainment, non-pension wealth, marital status, smoking, alcohol consumption, limiting illness, mobility impairment, and self-rated health, depressive symptoms, loneliness | Social isolation negatively associated with total activity count. Over the waking period (7am-10pm), social isolation positively associated with SB, negatively associated with LPA and MVPA. |
|  | None | None | None | No cross-sectional or longitudinal associations observed sleep (time spent in bed, TST, SOL) and loneliness. |

*Abbreviations*: LPA, light physical activity; MVPA, moderate-to-vigorous physical activity; SB, sedentary behaviour; SE, sleep efficiency; SOL, sleep onset latency; TST, total sleep time; UCLA, UCLA Loneliness Scale; WASO, Wake after sleep onset

**Table S6.** Findings of the articles using machine learning approaches

| **Study** | **Model performance** |
| --- | --- |
| Doryab et al., 2019 (USA)^21^ | Loneliness level (best model: all-epochs features):   - Accuracy: 80.2 - Precision: 80.3 - Recall: 80.1 - F1 score: 80.1   Change in loneliness (best model: semester-level features):   - Accuracy: 88.4 - Precision: 90.0 - Recall: 82.6 - F1 score: 81.0 |
| Jafarlou et al., (USA)^22^ | Smart ring data:   - Accuracy: 56.5 - Precision: 55.6 - Recall: 98.8 - F1 score: 71.1   Wrist-worn wearable data:   - Accuracy: 78.1 - Precision: 80.9 - Recall: 78.0 - F1 score: 79.4   Smartphone data:   - Accuracy: 81.0 - Precision: 87.7 - Recall: 75.6 - F1 score: 81.2   Smart ring + wrist-worn wearable + smartphone data:   - Accuracy: 82.2 - Precision: 88.3 - Recall: 77.3 - F1 score: 82.4 |
